# Malaria burden and care in hard-to-reach indigenous communities in the Peruvian Amazon during the COVID-19 pandemic: A mixed-methods study

**DOI:** 10.64898/2026.07.12.26357900

**Authors:** Susan Paredes Fernández, Luis M. Rojas, Joaquín Gómez, Julio Sandoval-Bances, Sandra Condori-Catachura, Paloma Diab García, Steven Abrams, Sibyl Anthierens, Hilde Bastiaens, Stella M. Chenet, Christopher Delgado-Ratto

**Affiliations:** Malaria Research group (MaRch), Global Health Institute, Department of Family Medicine and Population Health (FAMPOP), Faculty of Medicine, University of Antwerp, Antwerp, Belgium; Instituto de Investigación en Enfermedades Tropicales (IET), Universidad Nacional Toribio Rodríguez de Mendoza de Amazonas, Amazonas, Peru; Dirección Regional de Salud Amazonas, Amazonas, Peru; Research group Primary and Interdisciplinary care Antwerp (ELIZA), Department of Family Medicine and Population Health (FAMPOP), University of Antwerp, Antwerp, Belgium; Data Science Institute, Interuniversity Institute for Biostatistics and Statistical Bioinformatics, University of Hasselt, Diepenbeek, Belgium

**Author notes:** **Corresponding authors:** Prof. dr. Christopher Delgado-Ratto; Susan Paredes Fernandez, PhD Fellow.

**Keywords:** SARS-CoV-2, malaria, mixed-methods research, Amazon region, indigenous population, healthcare access

## Abstract

**Background:** The COVID-19 pandemic was associated with reported disruptions to healthcare delivery and malaria control activities in remote, malaria-endemic indigenous communities with limited access to healthcare. Remote indigenous communities in the Peruvian Amazon may have been especially vulnerable due to interruptions in malaria surveillance, diagnosis, and treatment. We investigated the impact of the COVID-19 pandemic on malaria burden, healthcare access, and community perceptions in hard-to-reach indigenous communities in the Amazonas region of Peru. In malaria-endemic regions, these lessons are particularly important because future health emergencies may similarly affect surveillance, diagnosis, prevention, and treatment activities, potentially reversing gains achieved through malaria control programs.

**Methods:** A convergent parallel mixed-methods study was conducted in 2021 and 2022 across four indigenous communities in the Río Santiago district, Amazonas region, Peru. Quantitative data were collected through cross-sectional malaria surveys, and qualitative data were obtained through semi-structured interviews with inhabitants, community health workers, and local health personnel. Malaria infections were detected by microscopy and confirmed by PCR. Factors associated with the occurrence of malaria infection were studied based on a generalized estimating equation (GEE) approach applied to clustered binary outcome data and accounting for repeated observations within participants across study years. Qualitative data were analyzed thematically, and both components were integrated using the Pillar Integration Process framework.

**Results:** A total of 514 participants were included. PCR-based malaria prevalence was 17.6% in 2021 and 26.09% in 2022. *P. falciparum* prevalence was particularly high in the Alianza Progreso community (2021: 24.19%, 2022: 16.16%) compared to the other studied communities. Previous malaria infection and chills were associated with PCR-confirmed malaria infection. Qualitative findings indicated that COVID-19-related disruptions affected malaria control activities by interrupting diagnosis and treatment availability, reducing community follow-up, and prioritizing COVID-19 response activities over malaria control. Participants also described fear of COVID-19, temporary migration to farm areas, and reliance on herbal medicine during the pandemic.

**Conclusions:** The COVID-19 pandemic may have contributed to exacerbated pre-existing challenges to malaria control in remote indigenous communities in the Peruvian Amazon. Strengthening malaria surveillance, ensuring continuity of diagnosis and treatment, and improving healthcare access in geographically isolated communities will be critical to supporting malaria elimination efforts in Peru.

**Author summary:** The COVID-19 pandemic disrupted essential health services worldwide, with potentially greater consequences for remote malaria-endemic communities with limited healthcare access. Indigenous populations in the Peruvian Amazon faced additional challenges due to interruptions in malaria surveillance, diagnosis, treatment, and control activities.

We assessed the impact of the COVID-19 pandemic on malaria burden and healthcare access in four remote indigenous communities in the Amazonas region of Peru by combining malaria screening with interviews involving community members, community health workers, and healthcare personnel. Our findings showed persistent malaria transmission during the pandemic period, particularly in communities affected by *Plasmodium falciparum*. Participants reported reduced access to malaria diagnosis and treatment, fewer community follow-up activities, fear of attending health facilities, and changes in healthcare-seeking behaviors.

These findings highlight how health emergencies can intensify existing barriers in geographically isolated populations and threaten malaria elimination efforts. Strengthening resilient health systems and ensuring continuity of essential malaria services during future crises will be critical to protect vulnerable communities in the Amazon region.

## Introduction

During the COVID-19 pandemic, major disruptions in healthcare delivery affected malaria control programs worldwide, particularly in low- and middle-income countries. Between 2020 and 2021, global malaria cases rose from 221 million to 241 million, and nearly 73% of malaria programs reported operational disruptions, with 19% experiencing severe interruptions in malaria-related services (1, 2). These disruptions contributed to an estimated 14 million additional malaria cases and 69,000 additional deaths in 2020 compared with pre-pandemic levels (1). Lockdowns, mobility restrictions, reduced access to healthcare, and fear of SARS-CoV-2 exposure further affected health-seeking behavior and the continuity of malaria diagnosis and treatment (3-5).

In Peru, the COVID-19 pandemic has exacerbated longstanding socioeconomic inequalities and major structural limitations in the healthcare system, particularly affecting remote and socially vulnerable populations in the Amazon region (1-3). During the pandemic, healthcare resources and public health priorities were largely redirected toward COVID-19 response activities, disrupting routine malaria surveillance, diagnosis, treatment, and vector control interventions. These challenges were especially pronounced in indigenous riverine communities, which are characterized by poor geographical accessibility, limited access to healthcare facilities, and persistent difficulties in sustaining malaria surveillance and control coverage (4).

Malaria transmission in Peru is concentrated in Amazonian regions, particularly Loreto, Amazonas, and Junín, which together account for approximately 98% of all reported malaria cases nationally (5). Transmission is highly heterogeneous and often sustained by asymptomatic and submicroscopic infections, complicating surveillance and elimination efforts in remote communities. Although intensified malaria control interventions substantially reduced the malaria burden in Loreto between 2017 and 2020 (reducing from 53178 to 13385 reported cases), the Amazonas region presented a particularly significant increase in reports during that period (increasing from 855 to 1553)(6, 7). Peru experienced a resurgence of malaria cases after the COVID-19 pandemic, with the number of cases rising from approximately 18,000 reported cases in 2019 to more than 27,000 in 2022 (8). This resurgence has been partially attributed to interruptions in malaria control activities during the pandemic (9).

In response to this epidemiological context, Peru launched a national malaria elimination plan in 2022, aiming to eliminate malaria by 2030 in priority areas and by 2045 nationwide (10). However, major challenges persist in remote indigenous communities, including limited access to healthcare, logistical barriers to diagnosis and treatment, a high proportion of undetected infections, and a situation which is further complicated by a limited understanding of how the COVID-19 pandemic may have affected malaria control efforts, therefore posing new threats to elimination efforts in the future when confronted with new emerging health threats (11, 12).

Although Amazonas was among the last regions in Peru to report COVID-19 cases, the virus rapidly spread throughout the northern province of Condorcanqui, home to the Awajún and Wampis indigenous populations. Interestingly, the mortality rate reported in Condorcanqui during 2020 (0.65%) was substantially lower than those death rates observed in the Amazonas region (1.47%) and Peru overall (3.7%)(11). Nevertheless, the broader consequences of the pandemic on healthcare access, disease surveillance, and malaria transmission in these remote communities remain poorly understood.

Despite the public health relevance of these populations and the overlapping challenges posed by malaria and COVID-19, no studies have simultaneously evaluated malaria burden, healthcare access, and community perceptions during the pandemic in remote Indigenous Amazonian settings. This knowledge gap limits the development of evidence-based strategies to strengthen malaria control and elimination efforts in these vulnerable populations.

Experiences during the COVID-19 pandemic provide valuable lessons for strengthening preparedness and response capacities for future public health emergencies and can reveal underlying weaknesses in health systems, particularly in remote and underserved settings. Therefore, we conducted a convergent parallel mixed-methods study in four hard-to-reach indigenous communities in the Río Santiago district of the Amazonas region in Peru. We combined cross-sectional malaria screening surveys with semi-structured interviews to investigate malaria prevalence, factors associated with infection, healthcare-seeking behavior, and perceptions of malaria prevention and care during the pandemic. By integrating epidemiological and qualitative evidence, we aimed to understand better how pandemic-related disruptions may have affected malaria burden and continuity of care in remote indigenous communities.

## Methods

### Study area and population

The study was conducted in four communities in the Río Santiago district, Condorcanqui province, in the Amazonas region of Peru: Alianza Progreso (AP), Caterpiza (CZ), Chapiza (CH), and Nueva Esperanza (NE), with data collected in October 2021 and between February and March 2022. The study communities are located along the Río Santiago River near the Ecuadorian border (Figure 1) and are accessible from Nieva town, the capital of Río Santiago, via river transport (>90 km).

**Figure 1.**
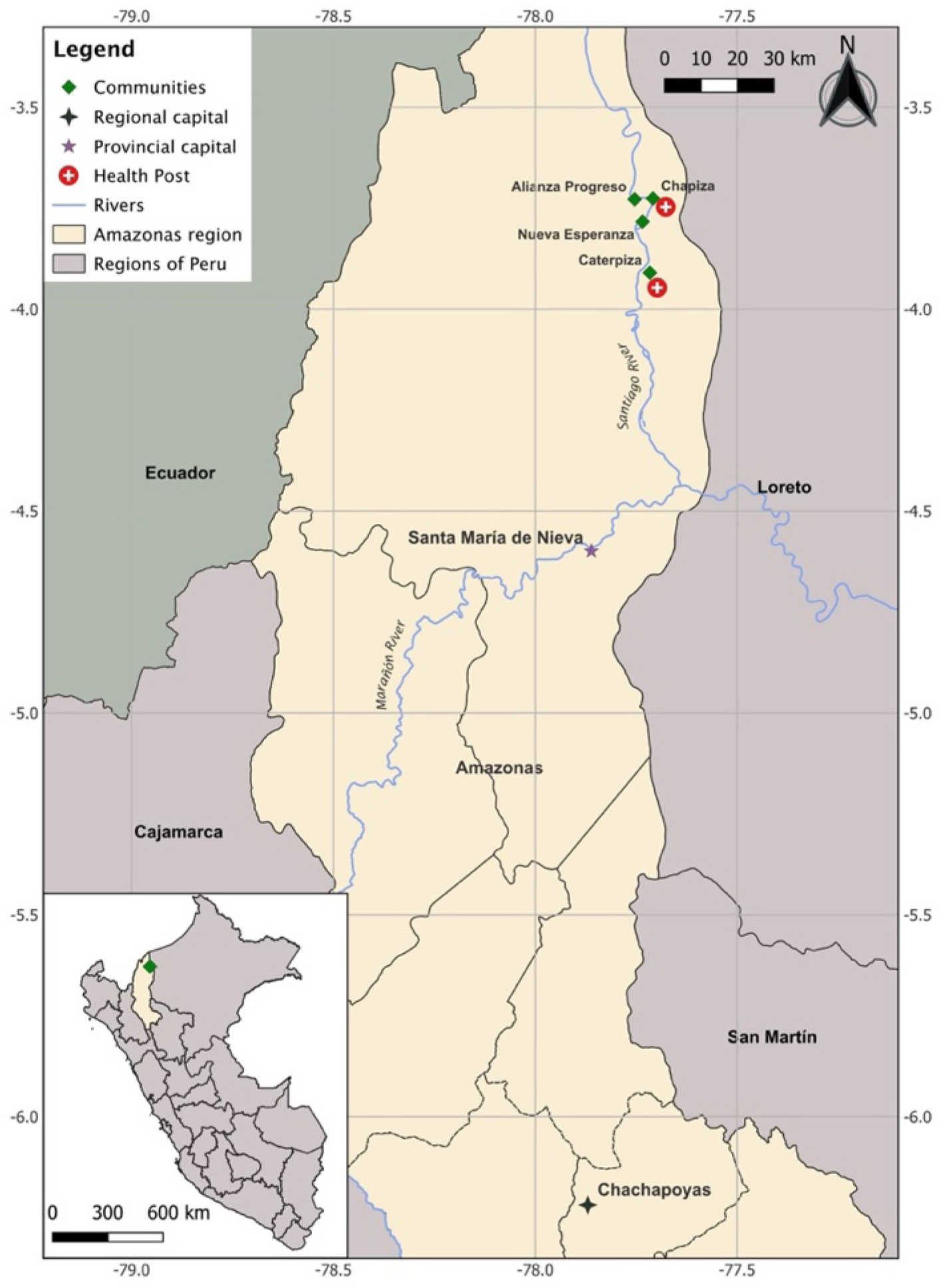
Location of the four study communities in the Río Santiago district, Alianza Progreso (AP), Caterpiza (CZ) Chapiza (CH) and Nueva Esperanza (NE), in Condorcanqui province, Amazonas region, Peru. The map was designed using QGIS Geographic Information System (Version 3.34.1), available at https://www.qgis.org. The data was visualized and placed in a spatial context using the OpenStreetMap base layer, available at https://www.openstreetmap.org. Regarding rivers, the geospatial information was obtained from the Peruvian national water authority (Autoridad Nacional del Agua - ANA), publicly available at https://snirh.ana.gob.pe/ConsultaIDE/.

The population is predominantly composed of the Awajún and Wampis indigenous groups living under conditions of socioeconomic vulnerability, with no access to potable water, electricity, and limited healthcare services. The Río Santiago district encompasses approximately 60 indigenous communities with a total population of 16,878 inhabitants and is located near the Ecuadorian border (13). Total inhabitants per study community were AP with 224 inhabitants, CZ with 414 inhabitants, CH with 950 inhabitants, and NE with 227 inhabitants.

Among the four study communities, only CH and CZ have health posts. Therefore, residents of AP and NE must travel by boat to access healthcare services in neighboring communities. In communities without health posts, individuals with suspected malaria are initially evaluated by community health workers (CHWs) trained by the Ministry of Health (MoH), who perform rapid diagnostic tests (RDTs) and support malaria prevention activities. Blood smears are then transported to the nearest health post for confirmation by light microscopy, the routine reference diagnostic method in the study area.

Treatment is administered according to the Peruvian national guidelines, based on *Plasmodium* species identification and adjusted for patient age and weight (14).

### Study design, sampling and data collection

A convergent parallel mixed-methods study with a cross-sectional design was conducted. Quantitative and qualitative data were collected concurrently, analyzed independently, and then integrated using the Pillar Integration Process (PIP) framework (15), which synthesizes both components by listing, matching, checking, and building pillars to generate a joint display of findings (Table 2).

Field activities were conducted in collaboration with personnel from the Dirección Regional de Salud (DIRESA) Amazonas (the local MoH dependency). In both 2021 and 2022, participant recruitment, sample collection, and data collection were conducted through active case detection (ACD). Recruitment strategies differed between study years: in 2021, participants were recruited through door-to-door household visits, whereas in 2022, recruitment was conducted through community-wide announcements using communal loudspeakers.

For the quantitative component, participants provided written informed consent before completing a structured questionnaire and undergoing malaria screening. Diagnostic samples included thick and thin blood smears for microscopy, which were examined by trained personnel from DIRESA Amazonas for routine malaria diagnosis and case management. Participants diagnosed with malaria received treatment according to national guidelines (16). In addition, dried blood spots collected on Whatman filter paper from all participants were transported to the Instituto de Investigación en Enfermedades Tropicales, Universidad Nacional Toribio Rodríguez de Mendoza (Amazonas, Peru), for molecular confirmation and further analysis.

For the qualitative component, semi-structured interviews were conducted and audio-recorded in February 2022. The semi-structured interviews explored perceptions and experiences related to malaria prevention, healthcare access, and malaria care during the COVID-19 pandemic. Separate interview guides in Spanish were developed for community members and healthcare personnel. Participants included inhabitants, health personnel, and community health workers, recruited through convenience sampling. This approach was considered appropriate given the remote location of the study settings, the limited number of healthcare workers involved in malaria, and the logistical challenges associated with accessing the community. Audio-recorded informed consent was obtained from all interviewees before data collection. Recruitment continued until the data sufficiently addressed the study’s scope (17).

Although Indigenous languages were the primary language spoken by participants in their communities, all interviewees were bilingual and demonstrated sufficient fluency in Spanish to participate comfortably in the interviews. Spanish was routinely used in healthcare interactions within the study setting and was therefore selected as the interview language. Participants were offered the opportunity to communicate in their preferred language; however, all interviews were conducted in Spanish, and the use of an interpreter was not necessary. This approach minimized potential translation-related data loss while facilitating direct communication between participants and the research team.

**Figure 2.**
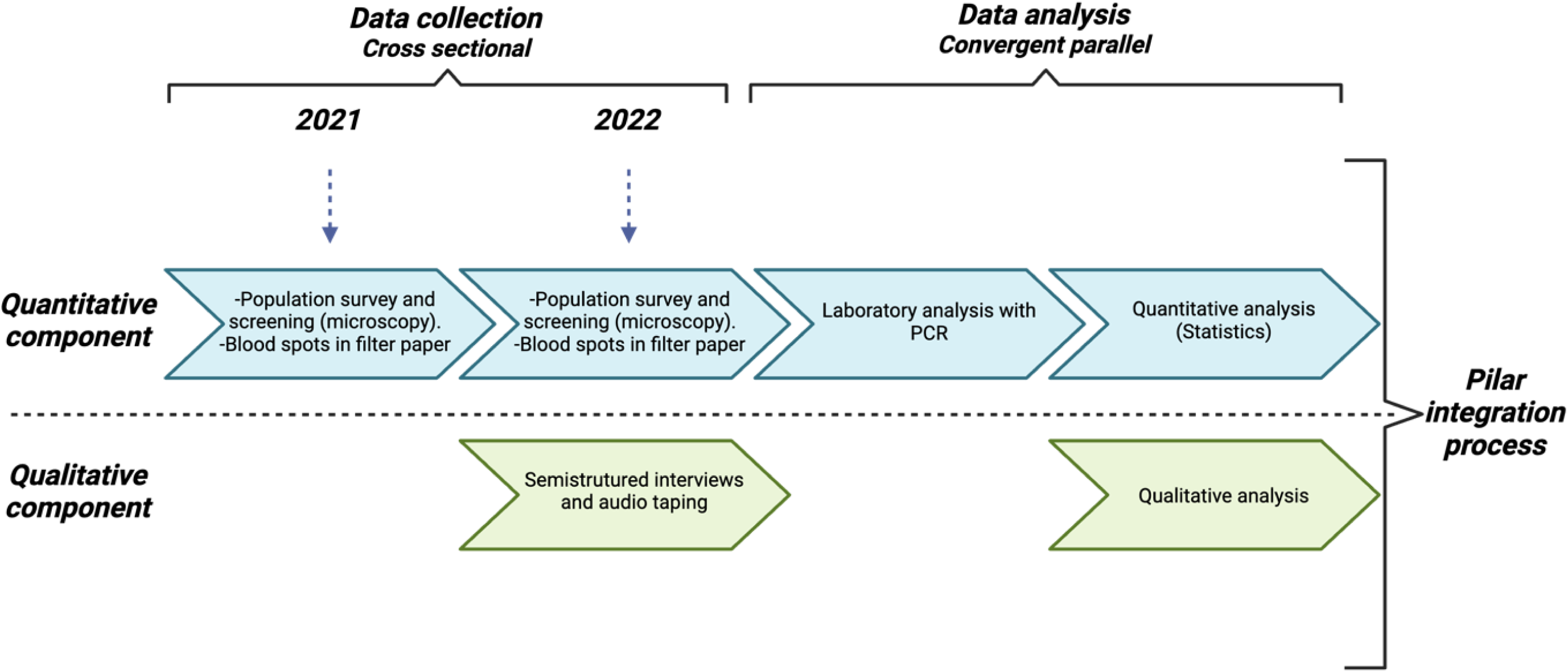
Overview of the convergent parallel mixed-methods study design, including quantitative and qualitative data collection, independent analyses, and integration via the Pillar Integration Process.

### Ethics

The study was approved by the Ethics Committee of the Universidad Nacional Toribio Rodríguez de Mendoza de Amazonas, Peru (approval code: CIEI-004). All adult participants (≥18 years) provided written informed consent before participating. For participants younger than 18 years, written informed consent was obtained from a parent or legal guardian, and assent was obtained from minors when appropriate.

### Laboratory procedures

Microscopy-based malaria diagnosis was conducted in the communities in accordance with Peruvian Ministry of Health (MoH) guidelines (14). Participants with microscopy-confirmed malaria infections received treatment from MoH personnel based on the identified *Plasmodium* species and national treatment guidelines. For quality control, a random subset of thick and thin blood smears was independently re-examined by an expert microscopist in the Laboratorio Intermedio de Salud Pública Condorcanqui.

In addition to blood smears, dried blood spots collected on Whatman filter paper were processed for molecular analysis. DNA was extracted from dried blood spots using the DNeasy Blood and Tissue Kit (17), and *Plasmodium* detection was performed using species-specific real-time PCR targeting the 18S rRNA gene, as previously described (18). Samples that were inconclusive by real-time PCR were subsequently confirmed by nested PCR (19).

All malaria infections detected by PCR were notified to the DIRESA Amazonas to ensure appropriate clinical management and follow-up in accordance with national malaria control procedures.

### Data analysis

Survey data were initially collected using paper-based forms and later digitized in REDCap 12.4.7. Quantitative data management and statistical analyses were conducted in R version 4.5.0 (R Core Team, 2025) using the *epiR* and *glmtoolbox* packages. Malaria prevalence and factors associated with infection were estimated based on PCR-confirmed infections.

Given PCR’s higher sensitivity compared with light microscopy, malaria infection was defined as a PCR-positive result for any Plasmodium species. Overall malaria prevalence was estimated and further stratified by Plasmodium species and study community. *Plasmodium spp*. prevalence values were compared across communities using Fisher’s exact test as some expected cell counts were too small. Because some participants were surveyed in both 2021 and 2022, a generalized estimating equation (GEE) approach was adopted to account for within-participant association induced through repeated measurements of the same individuals. We estimated two GEE models regressing the binary *Plasmodium spp.* PCR result against I) factors potentially influencing malaria prevalence and II) further including interaction effects to assess the possible effect modification by study year. Competing GEE models were compared using the Quasi-likelihood under the Independence Model Criterion (QICu), and the model with the lowest QICu value was selected as the best-fitting and most parsimonious model. All statistical tests were two-sided with a significance level of 0.05.

Interviews were audio-recorded, transcribed verbatim, and translated from Spanish to English by a trained, native Spanish-speaking member of the research team. Qualitative data were analyzed using thematic analysis, following the six-step framework of Braun and Clarke as described in the AMEE guideline (20, 21). Initial coding was conducted manually by one researcher, and codes, themes, and subthemes were subsequently reviewed and refined through iterative discussions among three members of the research team, including two epidemiologists (one conducted the interviews) and an MD with expertise in qualitative methods. The discussions facilitated the examination of alternative interpretations and helped ensure that emerging themes were grounded in participants’ accounts.

### Triangulation of results

In the mixed-methods analysis, methodological triangulation (quantitative and qualitative data) and data source triangulation (inhabitants, community health workers, and health personnel) were undertaken to examine convergence, complementarity, and discrepancies across findings. Integration was subsequently conducted using the PIP to construct a joint display (15). The process involved four stages: listing, matching, checking, and pillar building. Findings from both components were first analyzed separately and then compared across data sources and methods. Through an interactive and interpretative process, integrated themes (“pillars”) were generated to provide a comprehensive understanding (15).

## Results

### Sociodemographic and epidemiological characteristics

A total of 514 inhabitants were surveyed across the four communities. Individuals in CH were surveyed only in 2021, while residents in CZ were surveyed only in 2022. AP and NE inhabitants were surveyed in both study years.

The median participant age was 19 (IQR: 11 – 36), with AP and NE having a slightly older population than CH and CZ (Table 1). The proportion of participants without formal education was significantly different across communities (Annex 1, p-value < 0.001), with CZ (16.53%) and AP (16.15%) showing a higher observed value than CH (2.11%) and NE (8.03%) (Annex 1). The most frequently reported occupations were student (43.77%), farming (22.96%), and housewife (21.40%), while unemployment among adult participants was low (10.70%) (Annex 1).

**Table 1.**
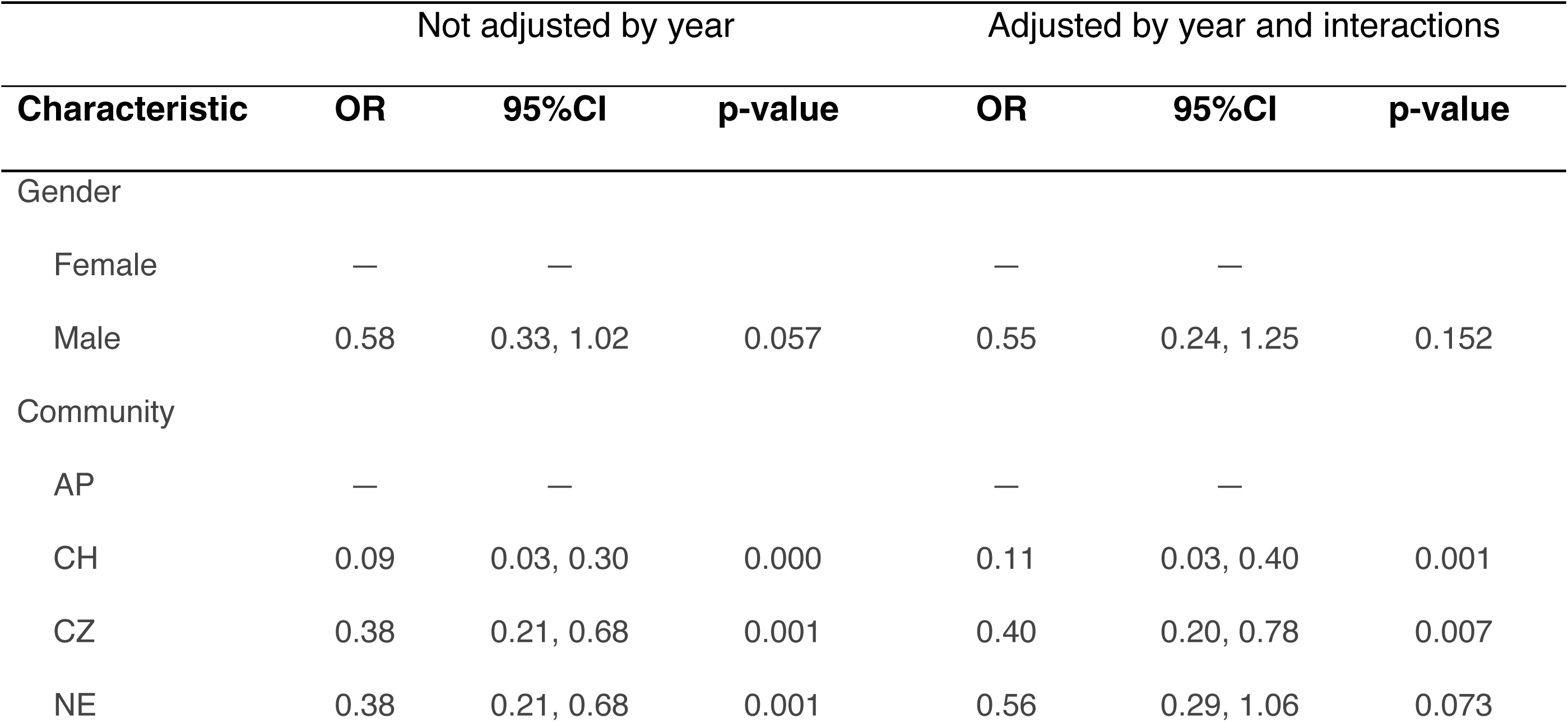
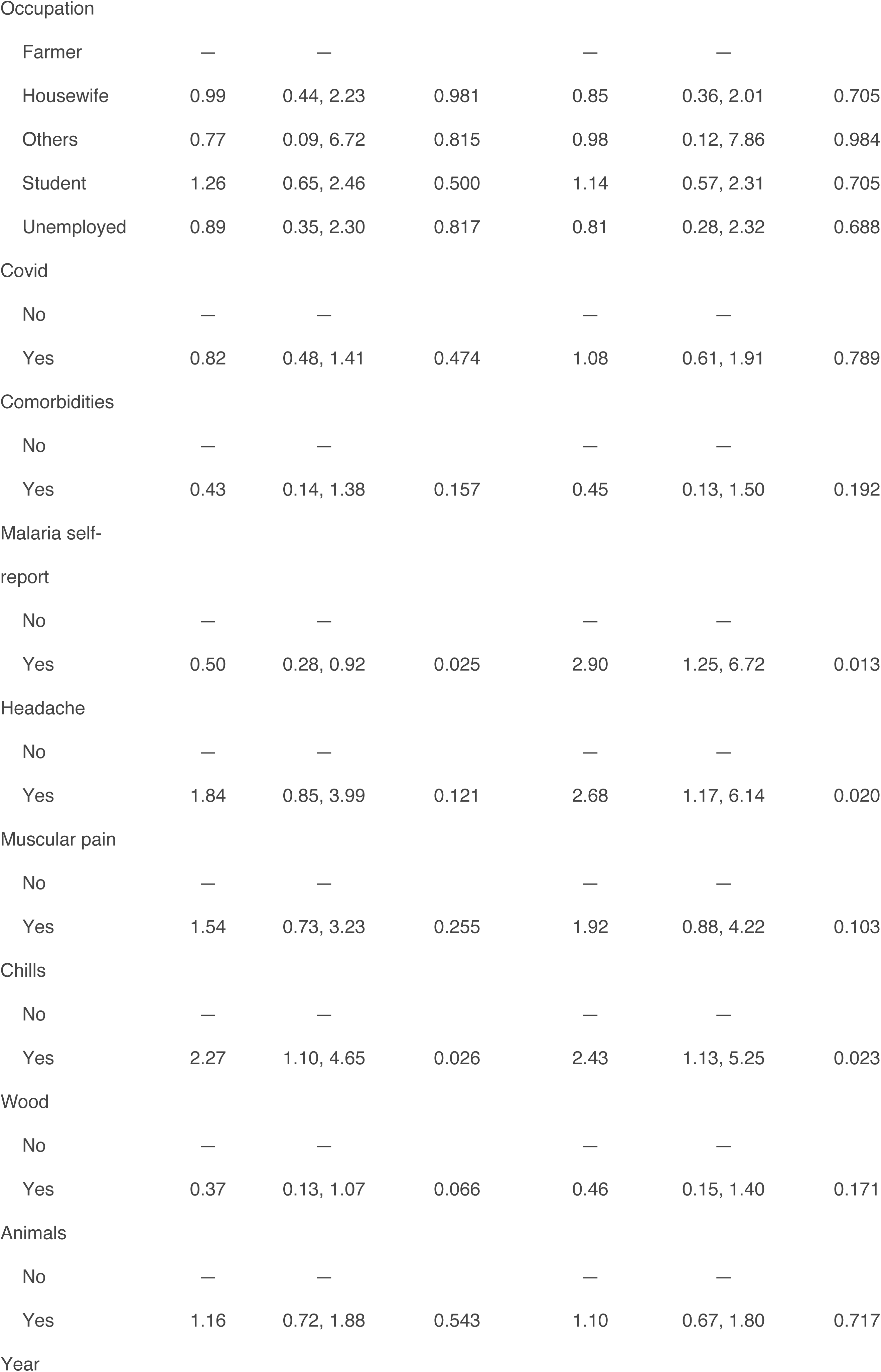
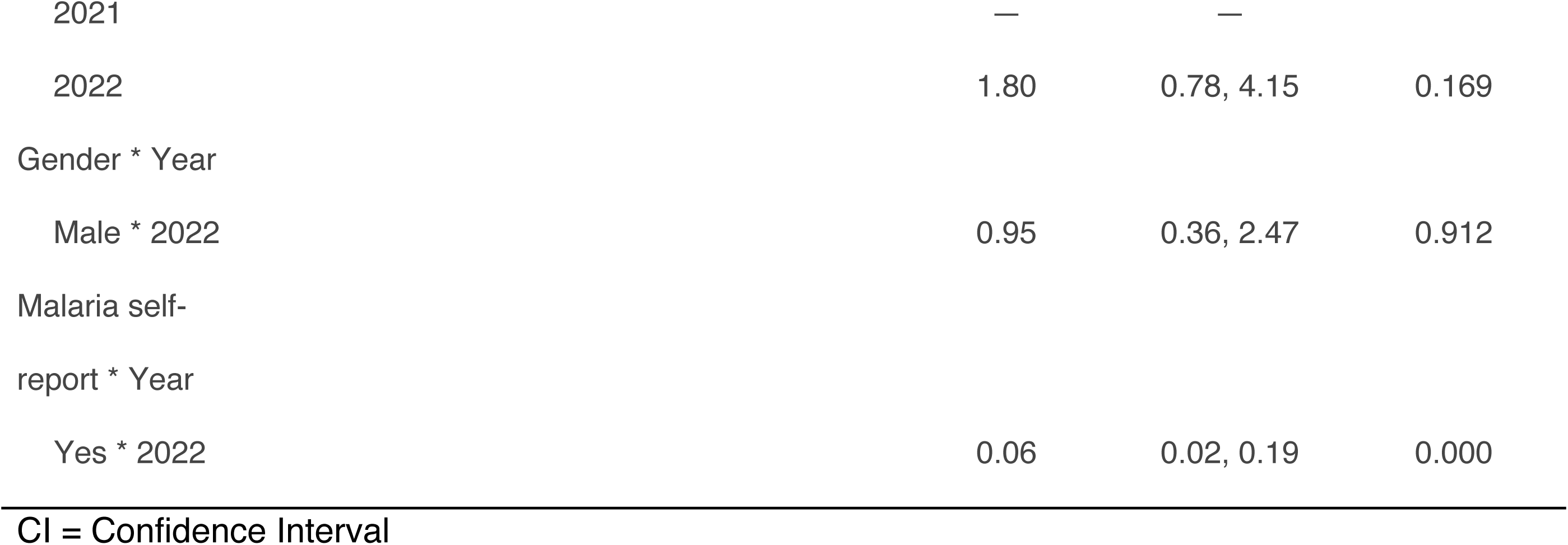
GEE analysis of factors associated with PCR-confirmed malaria infection.

**Table 2.**
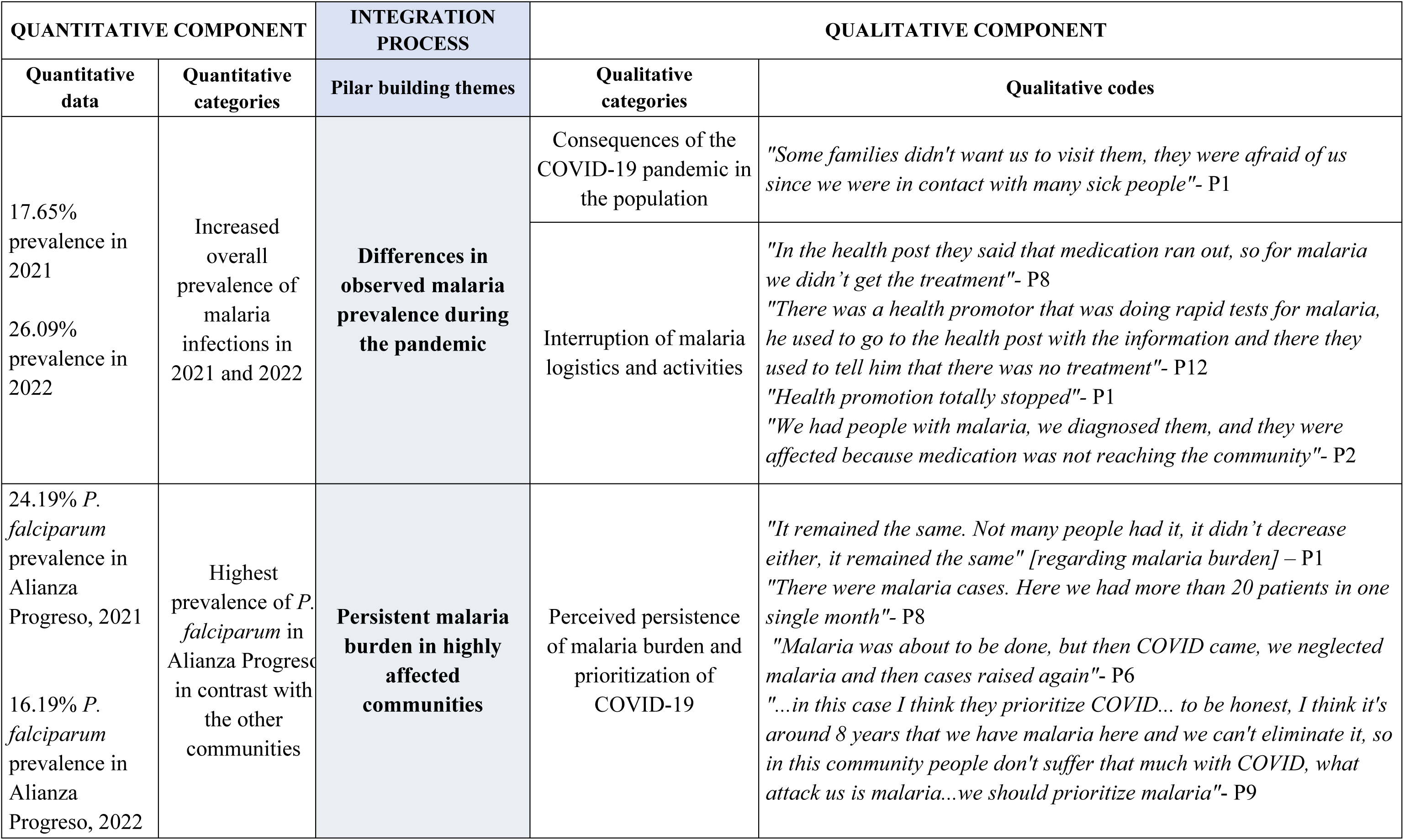
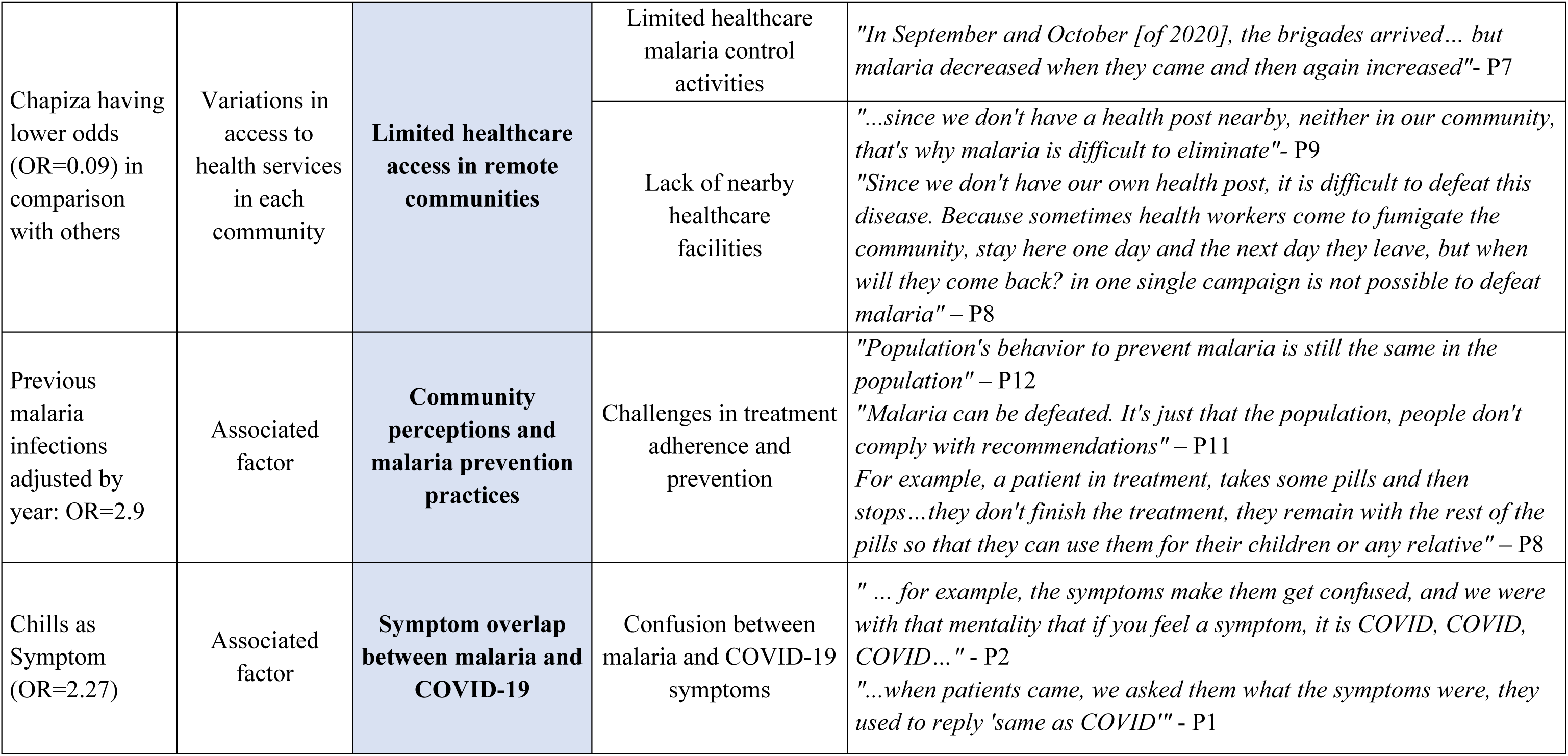
Joint display of integrated quantitative and qualitative findings.

| QUANTITATIVE COMPONENT |  | INTEGRATION PROCESS | QUALITATIVE COMPONENT |  |
| --- | --- | --- | --- | --- |
| Quantitative data | Quantitative categories | Pilar building themes | Qualitative categories | Qualitative codes |
| 17.65% prevalence in 2021<br>26.09% prevalence in 2022 | Increased overall prevalence of malaria infections in 2021 and 2022 | **Differences in observed malaria prevalence during the pandemic** | Consequences of the COVID-19 pandemic in the population | *"Some families didn't want us to visit them, they were afraid of us since we were in contact with many sick people"- P1* |
|  |  |  | Interruption of malaria logistics and activities | *"In the health post they said that medication ran out, so for malaria we didn't get the treatment"- P8*<br>*"There was a health promotor that was doing rapid tests for malaria, he used to go to the health post with the information and there they used to tell him that there was no treatment"- P12*<br>*"Health promotion totally stopped"- P1*<br>*"We had people with malaria, we diagnosed them, and they were affected because medication was not reaching the community"- P2* |
| 24.19% *P. falciparum* prevalence in Alianza Progreso, 2021<br>16.19% *P. falciparum* prevalence in Alianza Progreso, 2022 | Highest prevalence of *P. falciparum* in Alianza Progreso in contrast with the other communities | **Persistent malaria burden in highly affected communities** | Perceived persistence of malaria burden and prioritization of COVID-19 | *"It remained the same. Not many people had it, it didn't decrease either, it remained the same" [regarding malaria burden] – P1*<br>*"There were malaria cases. Here we had more than 20 patients in one single month"- P8*<br>*"Malaria was about to be done, but then COVID came, we neglected malaria and then cases raised again"- P6*<br>*"...in this case I think they prioritize COVID... to be honest, I think it's around 8 years that we have malaria here and we can't eliminate it, so in this community people don't suffer that much with COVID, what attack us is malaria...we should prioritize malaria"- P9* |
| Chapiza having lower odds (OR=0.09) in comparison with others | Variations in access to health services in each community | **Limited healthcare access in remote communities** | Limited healthcare malaria control activities | *"In September and October [of 2020], the brigades arrived... but malaria decreased when they came and then again increased"- P7* |
|  |  |  | Lack of nearby healthcare facilities | *"...since we don't have a health post nearby, neither in our community, that's why malaria is difficult to eliminate"- P9*<br>*"Since we don't have our own health post, it is difficult to defeat this disease. Because sometimes health workers come to fumigate the community, stay here one day and the next day they leave, but when will they come back? in one single campaign is not possible to defeat malaria" – P8* |
| Previous malaria infections adjusted by year: OR=2.9 | Associated factor | **Community perceptions and malaria prevention practices** | Challenges in treatment adherence and prevention | *"Population's behavior to prevent malaria is still the same in the population" – P12*<br>*"Malaria can be defeated. It's just that the population, people don't comply with recommendations" – P11*<br>*For example, a patient in treatment, takes some pills and then stops...they don't finish the treatment, they remain with the rest of the pills so that they can use them for their children or any relative" – P8* |
| Chills as Symptom (OR=2.27) | Associated factor | **Symptom overlap between malaria and COVID-19** | Confusion between malaria and COVID-19 symptoms | *" ... for example, the symptoms make them get confused, and we were with that mentality that if you feel a symptom, it is COVID, COVID, COVID..." - P2*<br>*"...when patients came, we asked them what the symptoms were, they used to reply 'same as COVID'" - P1* |

Use of bed nets as a malaria preventive measure varied substantially across communities, ranging from 27.27% in CZ to 83.85% in AP (Annex 1). Self-reported malaria episodes in the year preceding the survey were most frequent in AP, with half of the participants reporting a prior malaria infection and significant differences between communities (Annex 1).

Self-reported COVID-19 infections in the year preceding the survey were significantly higher in CZ (80.17%), followed by AP (55.90%), NE (37.96%), and CH (32.63%) (Annex 1).

### Results for quantitative analysis: malaria prevalence and associated factors

#### Malaria prevalence

Malaria prevalence by study year and community was estimated using both microscopy and PCR (Annex 2). Overall, PCR-based malaria was higher in 2022 (26.09%; 95% CI 21.01-31.69) than in 2021 (17.65%; 95% CI 13.02-23.10). In 2021, PCR malaria prevalence in AP (46.77%, 95% CI 33.98-59.88) was significantly different from the malaria prevalences in CH and NE (p-value<0.001, Fisher’s exact test), with lower observed prevalences in CH (6.32%, 95% CI 2.35-13.24) and NE (8.64%, 95% CI 3.55-17.0).

Divided by year and species, in 2021 AP showed a significantly different *P. vivax* (20.97%; 95% CI [11.66-33.18]) and *P. falciparum* (24.19%; 95% CI 14.22-36.74) PCR prevalence than other communities (Annex 4, p-value < 0.001), with higher observed prevalence values than CH and NE (Annex 4). A similar pattern was observed in 2022, with AP having significantly different *P. falciparum* prevalence than CZ and NE (Annex 4, p-value<0.05) and a higher observed prevalence (Annex 4). No significant differences were observed in *P. vivax* prevalence between communities. Notably, the highest *P. vivax* prevalence was found in NE (28.57%; 95% CI 17.3-42.21) (Annex 4).

#### Factors associated with malaria infection

The mean model, including the variables gender, community, occupation, COVID self-report, comorbidities, previous malaria self-report, headache, muscular pain, chills, wood house materials, and animals surrounding the households, showed the best fit compared with the full model (full model QICu: 442.64 vs final model QICu: 423.153). Interactions with the study year were further evaluated to assess whether associations differed between 2021 and 2022 (QICu: 372.39) (Table 1).

When we do not adjust by year, participants reporting previous malaria infections had lower odds of PCR-confirmed *Plasmodium spp.* infection (OR: 0.5, 95% CI 0.28-0.92) while adjusting by other factors (Table 1). Similarly, the presence of chills during screening was strongly associated with PCR-confirmed *Plasmodium spp.* (OR: 2.27, 95%CI 1.10-4.65). In contrast, residing in CH (0.09, 95% CI 0.03-0.30), CZ (0.38, 95% CI 0.21-0.68), or NE (0.38, 95% CI 0.21-0.68) was associated with lower odds of malaria infection compared with living in AP, after adjusting for covariates (Table 1).

For the GEE approach, including year and interactions between malaria self-report, gender and year revealed an effect modification between year and malaria self-report. In 2021, participants who self-reported a previous malaria infection had 2.90 (95%CI 1.25-6.72) times higher odds of a PCR-confirmed *Plasmodium spp.* infection compared to those who did not. However, this association was reversed by 2022, indicating that the relationship between malaria self-report and current PCR-confirmed infection was not consistent across study years (Table 1).

### Results of qualitative analysis: thematic analysis of semi-structured questionnaires

#### Perception of the COVID-19 pandemic on malaria care and burden

Fourteen semi-structured interviews, each lasting 10 to 30 minutes, were conducted. Interviewees included three local MoH health personnel (one female and two males), two community health workers (one female and one male), and nine community inhabitants (two females and seven males). To avoid deductive disclosure, participants were identified by participant codes P1 to P14.

Thematic analysis identified three major themes and eight subthemes regarding the impact of the COVID-19 pandemic on malaria prevention, diagnosis, and healthcare access in the study communities.

#### Community impacts of the COVID-19 pandemic

Participants described major disruptions in social, economic, educational, and healthcare systems during the COVID-19 pandemic: *“COVID has affected a lot in the economy, politics and socially”* [P8]. Interviewees also highlighted the central role of healthcare personnel in COVID-19 response activities, including community-level monitoring and case follow-up, supported by internal strategies such as the “purple alert”, which was implemented by the health personnel to control the re-emergence of COVID-19 cases.

#### Malaria care disruptions during the COVID-19 pandemic

Participants reported substantial interruptions to malaria control activities during the pandemic, including reduced surveillance, medication shortages, and delays in malaria diagnosis and treatment: *“There was a community health promoter that was doing rapid tests for malaria; he used to go to the health post with the information, and there they used to tell him that there was no treatment”*[P3].

Several interviewees perceived that healthcare efforts were prioritized for COVID-19 response activities over malaria control: *“… in this case, they prioritized COVID… to be honest, I think it’s around 8 years that we have malaria here, and we can’t eliminate it, so in this community people don’t suffer that much with COVID, what attacks us is malaria…we should prioritize malaria”* [P8]

#### Barriers to malaria diagnosis and malaria prevention behavior in the community

Although malaria-related activities gradually resumed after 2020, participants emphasized persistent barriers to healthcare access, especially in communities without health posts, and highlighted that population behavior towards prevention is weak: *“Malaria can be defeated. It’s just that the population, people don’t comply with recommendations”* [P5]. Given the shortage of malaria treatment, interviewees also reported relying on herbal or traditional medicine, and also did the same practice for COVID-19: *“Most people in the jungle are cured with herbal medicine”* [P8].

### Results of the joint display with quantitative and qualitative components

Quantitative and qualitative findings were integrated using a joint display within the PIP framework. Five integrated themes were built up from the combined analysis, highlighting how COVID-19-related disruptions affected malaria burden, healthcare access, continuity of malaria control activities, and community perceptions of malaria prevention and treatment (Table 3).

### Differences in observed malaria prevalence during the pandemic

The increased malaria prevalence observed during the COVID-19 pandemic appears to reflect not only ongoing transmission but also the indirect effect of health system disruption, whereby interruptions in malaria surveillance, diagnosis, prevention, and treatment activities may have reduced the capacity to detect and control infections in remote communities.

### Persistent malaria burden in highly affected communities

The convergence of epidemiological data and community and health workers’ experiences suggests that malaria has become a normalized and enduring component of daily life in highly endemic communities, reflecting persistent transmission despite ongoing control efforts.

### Limited healthcare access in remote communities

Geographical isolation emerges as a fundamental determinant of malaria risk, linking limited access to healthcare services, reducing treatment opportunities and continue community-level transmission, thereby reinforcing health inequities in remote Amazonian settings.

### Community perceptions and malaria prevention practices

The persistence of malaria among individuals with previous infection histories suggests that the experience with the disease does not necessarily translate into effective prevention, highlighting how structural and behavioral barriers may limit the adoption and sustainability of preventive practices.

### Symptom overlap between malaria and COVID-19

The overlap between malaria and COVID-19 symptomatology may have introduced diagnostic uncertainty during the pandemic, potentially affecting symptom recognition and timely case management, thereby masking the true burden of malaria in remote communities.

## Discussion

The present study identified a substantial malaria burden in remote indigenous communities from the Amazonas region during the COVID-19 pandemic. Moreover, the prevalence estimates obtained from PCR and microscopy underscore the presence of submicroscopic infections that remain undetected in routine surveillance, highlighting the need to strengthen diagnostic approaches by deploying molecular tools in such settings. Similar findings have been reported in previous studies from Peru and other endemic settings, where molecular diagnosis improved detection of asymptomatic and low-density infections that are frequently missed by microscopy (22-25). Likewise, a recent report from indigenous communities in Amazonas revealed that nearly 75% of submicroscopic infections were asymptomatic (12). These findings are particularly relevant within the framework of the Peruvian National Malaria Elimination Plan 2022-2030.

Although malaria prevalence was estimated to be higher in 2022 than in 2021, these differences should be interpreted with caution because participant recruitment strategies differed between study years. Door-to-door household recruitment was used in 2021, whereas opportunistic recruitment through community announcements was implemented in 2022, potentially increasing participation of symptomatic individuals.

Our findings suggest that malaria control activities were affected during the COVID-19 pandemic in the studied communities. Quantitative and qualitative findings jointly indicated interruptions in malaria diagnosis, treatment availability, health promotion activities, and community follow-up. Interviewees reported medication shortages, interruptions to malaria surveillance activities, and the prioritization of COVID-19 response efforts over malaria control. Similar disruptions in malaria care services during the pandemic have been reported in several low-and middle-income countries, including diagnosis, treatment, and supply chains (26-30).

Moreover, respondents were aware of how the pandemic has affected them. Most interviewees highlighted how specific aspects of their lives (economic, social, and political) were affected: *“What I can say is that COVID-19 has affected a lot in the economy, politics, and socially”* [P8]. These consequences were also identified by other studies in rural areas (26, 31), they reported that poor households were adversely affected, leading to not only social and economic imbalance but also to potential psychological issues due to stress. Although, mental health issues were not mentioned or clearly identified by our interviewees, COVID-19 certainly caused alert and fear in the population: *"A lot of inhabitants have suffered due to this disease, but more they were feared to COVID-19"* [P2].

Communities without health posts faced greater barriers to accessing healthcare and malaria follow-up. Participants emphasized that malaria control activities often relied on periodic visits from healthcare brigades (for screening and treatment), and access to diagnosis and treatment was abruptly interrupted and complicated to perform during the pandemic: *"All the health attention, planning (family planning), control, immunization, and schools were stopped"* [P13]; *“There was a community health promotor that was doing rapid tests for malaria, he used to go to the health post with the information and there they used to tell him that there was no treatment”* [P3]*; “The medication was not reaching the communities”* [P1]. These limitations are consistent with the high malaria prevalence observed in some communities, particularly in Alianza Progreso and Nueva Esperanza. Together, these findings highlight the vulnerability of remote indigenous communities to interruptions in healthcare delivery during public health emergencies.

Although *P. vivax* is reported as the predominant malaria species in Peru, our results showed a high prevalence of *P. falciparum* in Alianza Progreso during both study years. Previous reports from Loreto and Amazonas have documented increasing *P. falciparum* transmission and re-emergence in areas where the species had previously declined (24, 25, 32, 33). The persistence of *P. falciparum* transmission in these remote communities demands further epidemiological and molecular investigation, particularly given its implications for malaria elimination efforts. Moreover, such variations in both years could also be partially explained by the strong seasonality of malaria transmission, since the first intervention was conducted during dry season (October 2021) and the second during rainy season (February 2022) (34).

Previous malaria infections were associated with higher odds of PCR-confirmed malaria infection, which may reflect persistent exposure to malaria transmission or recurrent infections. Additionally, the qualitative results show a discrepancy in perceptions of the malaria burden in the community during the pandemic. Interviewed community members perceived malaria to be highly prevalent within their communities: “*There were malaria cases. Here we had more than 20 patients in one single month”* [P3]; however, they were unable to determine whether the frequency had increased compared with the pre-pandemic period. This perception may be influenced by linguistic and cultural factors, as the term used for malaria in their local indigenous language is synonymous with “fever.” Consequently, febrile illnesses may be more readily interpreted as malaria, potentially shaping community perceptions of the disease occurrence and burden. On the other hand, healthcare personnel perceived malaria incidence as relatively stable during the pandemic: *“It remained the same. Not many people had it, it didn’t decrease either, it remained the same”* [P1]; however, they acknowledged limited awareness of cases in communities accessible for routine visits and surveillance activities. This discrepancy may reflect differences in how malaria burden is experienced and measured. Community perceptions are grounded in lived experience and repeated exposure to illness within households and communities, whereas health personnel rely primarily on reported cases and routine surveillance data. In geographically isolated settings, where barriers to healthcare access may limit case detection, community experiences may capture dimensions of disease burden that remain partially invisible to formal health information systems. The finding highlights the importance of integrating community perspectives with epidemiological surveillance to achieve a more comprehensive understanding of malaria dynamics in hard-to-reach populations.

Regarding presenting clinical symptoms, chills were also strongly associated with malaria infections. However, interviewees reported symptom overlap between malaria and COVID-19, potentially complicating community-level recognition and health-seeking behavior during the pandemic due to similar symptomatology.

Our qualitative findings revealed that the population in these communities feared COVID-19, altering their health-seeking behavior and healthcare access. Interviewees reportedly abandoned their homes and temporarily moved to distant farm areas to avoid contact with other community members: *“They were in other places; this means that some of them were not in the community. They ran away for 3, 4 or 5 hours-walk to their farm plots. We couldn’t do a follow-up for those who were with malaria”* [P2]. This behavior limited follow-up by healthcare personnel, potentially affecting malaria diagnosis and treatment continuity. Moreover, interviewees reported relying on herbal or traditional medicine to manage malaria and COVID-19, reflecting adaptive healthcare practices in remote settings with limited healthcare access.

Overall, our results show that the COVID-19 pandemic may have worsened existing difficulties in malaria control among remote indigenous communities in the Peruvian Amazon. Enhancing malaria surveillance, maintaining consistent diagnosis and treatment, and expanding healthcare access in these geographically isolated areas are crucial steps to support Peru’s efforts to eliminate malaria.

### Strengths and limitations

This study has several strengths. First, the convergent parallel mixed-methods design with formal integration via the Pillar Integration Process allowed quantitative and qualitative findings to be systematically combined, generating insights that neither component could have produced alone. Second, malaria diagnosis was confirmed by PCR, enabling detection of submicroscopic infections that routine microscopy misses, a meaningful methodological advantage in a setting where asymptomatic and low-density infections are known to be prevalent. Third, the study was conducted in genuinely hard-to-reach indigenous communities in the Río Santiago district, a population that is both epidemiologically relevant to Peru’s elimination agenda and substantially underrepresented in the malaria literature. Fourth, data source triangulation across community members, community health workers, and health personnel provided multiple perspectives on the same phenomena.

Several limitations should be acknowledged. First, the cross-sectional design only allowed estimation of malaria prevalence for each year and precluded the establishment of a temporal sequence between risk factors and malaria infections. Second, the two study rounds are not directly comparable: door-to-door recruitment in 2021 and community-wide announcement-based recruitment in 2022 introduce different participation biases, most plausibly an overrepresentation of symptomatic individuals in 2022, which may partly explain the higher observed prevalence that year. Third, the convenience sample of 14 interviews, with predominantly male community participants, may not fully capture the range of community experiences, particularly regarding women’s health-seeking behavior and prevention practices.

## Conclusions

The COVID-19 pandemic may have exacerbated pre-existing challenges to malaria control in remote indigenous communities in the Peruvian Amazon. The combined quantitative and qualitative findings suggest that disruptions in healthcare delivery, interruptions in malaria control activities, limited access to health services, and persistent barriers to community follow-up may have affected malaria surveillance, diagnosis, and continuity of treatment during the pandemic.

Our findings highlight the vulnerability of geographically isolated indigenous communities to disruptions in essential healthcare services during public health emergencies. Strengthening malaria surveillance, improving access to diagnosis and treatment, and ensuring continuity of malaria interventions in remote settings will be critical to supporting malaria elimination efforts in Peru.

## Data Availability

The dataset is available upon request to the corresponding author, provided the ethics committee approves data sharing. An overview of the data results is presented in the Supplementary files.

## Acknowledgements

We thank all study participants from the study communities who agreed to take part in the project, especially those who participated in the interviews, for trusting us to delve into their perspectives and opinions on malaria in a context as challenging as the COVID-19 pandemic. We also want to thank the field team from the IET- UNTRM and DIRESA Amazonas for sample and data collection, and quality control, respectively.

This work was primarily supported by the Global Minds Small Research Grant 2021 project entitled “Evaluating the impact of COVID-19 pandemic on the malaria burden in indigenous communities of the Peruvian Amazon”, and the FONDECYT project “Metagenomics in vectors of the Amazon region: Identifying potential hotspots for emerging and re-emerging diseases” (Project Contract N°050-2021-FONDECYT). Additional support was provided through the VLIRUOS ICP Master of Epidemiology (MEPI) programme and VLIRUOS TEAM project “Strengthening national efforts to eliminate malaria in Peru (CeroMalariaPeru)

## Authors’ contribution

Study conception: CDR, HB, SA, PDG

Study design: CDR, HB, SA, PDG

Field work and supervision: SPF, LMR, SMC

Laboratory work: LMR, JSB

Data analysis and interpretation: SPF, JG, SA, HB, CDR

First draft: SPF, SCC, CDR

Review manuscript draft: All authors

All authors read and approved the final manuscript.

## References

1. Díaz-Ruiz R, Vargas-Fernández R, Rojas-Roque C, Hernández-Vásquez A. Socioeconomic inequalities in the use of medical consultation services in Peru, 2019. International Journal for Equity in Health. 2024;23(1).

2. Schwalb A, Seas C. The COVID-19 Pandemic in Peru: What Went Wrong? The American Journal of Tropical Medicine and Hygiene. 2021;104(4):1176–8.

3. Flores López MG, Soto Tarazona A, De La Cruz-Vargas JA. Regional distribution of COVID-19 mortality in Peru. Revista de la Facultad de Medicina Humana. 2021;21(2):326–34.

4. Carrasco-Escobar G, Villa D, Barja A, Lowe R, Llanos-Cuentas A, Benmarhnia T. The role of connectivity on malaria dynamics across areas with contrasting control coverage in the Peruvian Amazon. PLOS Neglected Tropical Diseases. 2024;18(11):e0012560.

5. Ministerio de Salud Perú. Número de casos de malaria, Perú 2023-2024. Ministerio de Salud, Perú; 2024.

6. Ministerio de Salud P. Plan Malaria Cero 2017-2021. 2017.

7. Ministerio de Salud Perú. Número de casos de malaria Perú 2015-2021. 2021:14.

8. Ministerio de Salud P. Alerta Epidemiológica: Incremento de casos y ocurrencia de brotes por malaria a nivel nacional. 2022. p. 3.

9. Hogan AB, Jewell BL, Sherrard-Smith E, Vesga JF, Watson OJ, Whittaker C, et al. Potential impact of the COVID-19 pandemic on HIV, tuberculosis, and malaria in low-income and middle-income countries: a modelling study. The Lancet Global Health. 2020;8(9):e1132–e41.

10. Ministerio de Salud P. Documento técnico: Plan hacia la eliminación de la malaria en el Perú 2022-2030. In: Lima, editor. Lima, Peru: Ministerio de Salud, Perú; 2022.

11. Pajuelo-Reyes C, Rojas LM, Campos CJ, Saavedra-Samillan M, Bernal JM, Tejedo JR, et al. Malaria and COVID-19 in native communities of Amazonas, Peru. Revista de la Facultad de Medicina Humana. 2022;22(3):533–9.

12. Saavedra-Samillán M, López-Ugarte LC, Sandoval-Bances J, Rojas LM, Udhayakumar V, Valdivia HO, et al. Hidden burden: submicroscopic Plasmodium spp. infections in indigenous populations of the Peruvian Amazon. Malaria Journal. 2025;25(1).

13. Instituto Nacional de Estadística e Informática P. Resultados definitivos de los censos nacionales 2017 - Amazonas - Tomo 01. Instituto Nacional de Estadística e Informática, Perú; 2018.

14. Ministerio de Salud P. Norma técnica de salud para la atención de la malaria y malaria grave en el Perú. 2015.

15. Johnson RE, Grove AL, Clarke A. Pillar Integration Process: A Joint Display Technique to Integrate Data in Mixed Methods Research. Journal of Mixed Methods Research. 2019;13(3):301–20.

16. Ministerio de Salud P. Norma técnica de salud para la atención integral de malaria no complicada y malaria grave en el Perú. 2025.

17. Malterud K, Siersma VD, Guassora AD. Sample Size in Qualitative Interview Studies. Qualitative Health Research. 2016;26(13):1753–60.

18. Rougemont M, Van Saanen M, Sahli R, Hinrikson HP, Bille J, Jaton K. Detection of Four Plasmodium Species in Blood from Humans by 18S rRNA Gene Subunit-Based and Species-Specific Real-Time PCR Assays. Journal of Clinical Microbiology. 2004;42(12):5636–43.

19. Snounou G, Singh B. Nested PCR Analysis of Plasmodium Parasites. Humana Press; 2002. p. 189-204.

20. Braun V, Clarke V. Using thematic analysis in psychology. Qualitative Research in Psychology. 2006;3(2):77–101.

21. Kiger ME, Varpio L. Thematic analysis of qualitative data: AMEE Guide No. 131. Medical Teacher. 2020;42(8):846-54.

22. Gholizadeh S, Naseri Karimi N, Zakeri S, Dinparast Djadid N. The Role of Molecular Techniques on Malaria Control and Elimination Programs in Iran: A Review Article. Iran J Parasitol. 2018;13(2):161–71.

23. Baldeviano GC, Okoth SA, Arrospide N, Gonzalez RV, Sánchez JF, Macedo S, et al. Molecular Epidemiology of Plasmodium falciparum Malaria Outbreak, Tumbes, Peru, 2010–2012. Emerging Infectious Diseases. 2015;21(5).

24. Kattenberg JH, Fernandez-Miñope C, Van Dijk NJ, Llacsahuanga Allcca L, Guetens P, Valdivia HO, et al. Malaria Molecular Surveillance in the Peruvian Amazon with a Novel Highly Multiplexed Plasmodium falciparum AmpliSeq Assay. Microbiology Spectrum. 2023;11(2).

25. Montenegro CC, Bustamante-Chauca TP, Pajuelo Reyes C, Bernal M, Gonzales L, Tapia-Limonchi R, et al. Plasmodium falciparum outbreak in native communities of Condorcanqui, Amazonas, Perú. Malaria Journal. 2021;20(1).

26. Shafi M, Liu J, Jian D, Rahman IU, Chen X. Impact of the COVID-19 pandemic on rural communities: a cross-sectional study in the Sichuan Province of China. BMJ Open. 2021;11(8):e046745.

27. Singh DR, Sunuwar DR, Shah SK, Karki K, Sah LK, Adhikari B, et al. Impact of COVID-19 on health services utilization in Province-2 of Nepal: a qualitative study among community members and stakeholders. BMC Health Services Research. 2021;21(1).

28. Zawawi A, Alghanmi M, Alsaady I, Gattan H, Zakai H, Couper K. The impact of COVID-19 pandemic on malaria elimination. Parasite Epidemiology and Control. 2020;11:e00187.

29. Torres K, Alava F, Soto-Calle V, Llanos-Cuentas A, Rodriguez H, Llacsahuanga L, et al. Malaria Situation in the Peruvian Amazon during the COVID-19 Pandemic. Am J Trop Med Hyg. 2020;103(5):1773–6.

30. Aborode AT, David KB, Uwishema O, Nathaniel AL, Imisioluwa JO, Onigbinde SB, et al. Fighting COVID- 19 at the Expense of Malaria in Africa: The Consequences and Policy Options. The American Journal of Tropical Medicine and Hygiene. 2021;104(1):26–9.

31. Zhang Y, Ma ZF. Impact of the COVID-19 Pandemic on Mental Health and Quality of Life among Local Residents in Liaoning Province, China: A Cross-Sectional Study. International Journal of Environmental Research and Public Health. 2020;17(7):2381.

32. Fernandez-Miñope C, Delgado-Ratto C, Contreras-Mancilla J, Ferrucci HR, Llanos-Cuentas A, Gamboa D, et al. Towards one standard treatment for uncomplicated Plasmodium falciparum and Plasmodium vivax malaria: Perspectives from and for the Peruvian Amazon. International Journal of Infectious Diseases. 2021;105:293–7.

33. Carrasco-Escobar G, Miranda-Alban J, Fernandez-Miñope C, Brouwer KC, Torres K, Calderon M, et al. High prevalence of very-low Plasmodium falciparum and Plasmodium vivax parasitaemia carriers in the Peruvian Amazon: insights into local and occupational mobility-related transmission. Malar J. 2017;16(1):415.

34. Angel Rosas-Aguirre NS, Alejandro Llanos-Cuentas, Anna Rosanas-Urgell, Gabriel Carrasco-Escobar, Hugo Rodriguez, Dionicia Gamboa, Juan Contreras-Mancilla, Freddy Alava, Irene S. Soares, Edmond Remarque, Umberto D’Alessandro, Annette Erhart. Hotspots of Malaria Transmission in the Peruvian Amazon: Rapid Assessment through a Parasitological and Serological Survey. PLoS ONE. 2015.

